# Novel polygenic risk score links depression-related cortical transcriptomic changes to brain morphology and symptom severity

**DOI:** 10.1101/2021.02.28.21251786

**Authors:** Amy E Miles, Fernanda C Dos Santos, Enda M Byrne, Miguel E Renteria, Andrew M McIntosh, Mark J Adams, Giorgio Pistis, Enrique Castelao, Martin Preisig, Bernhard T Baune, K Oliver Schubert, Cathryn M Lewis, Lisa A Jones, Ian Jones, Rudolf Uher, Jordan W Smoller, Roy H Perlis, Douglas F Levinson, James B Potash, Myrna M Weissman, Jianxin Shi, Glyn Lewis, Brenda WJH Penninx, Dorret I Boomsma, Steven P Hamilton, Major Depressive Disorder Working Group of the Psychiatric Genomics Consortium, Etienne Sibille, Ahmad R Hariri, Yuliya S Nikolova

## Abstract

Our group developed a transcriptome-based polygenic risk score (T-PRS) that uses common genetic variants to capture ‘depression-like’ shifts in cortical gene expression. Here, we mapped T-PRS onto diagnosis and symptom severity in major depressive disorder (MDD) cases and controls from the Psychiatric Genomics Consortium (PGC). To evaluate potential mechanisms, we further mapped T-PRS onto discrete measures of brain morphology and broad depression risk in healthy young adults. Genetic, self-report, and/or neuroimaging data were available in 29,340 PGC participants (59% women; 12,923 MDD cases, 16,417 controls) and 482 participants in the Duke Neurogenetics Study (DNS: 53% women; aged 19.8±1.2 years). T-PRS was computed from SNP data using PrediXcan to impute cortical expression levels of MDD-related genes from a previous post-mortem transcriptome meta-analysis. Sex-specific regressions were used to test effects of T-PRS on depression diagnosis, symptom severity, and Freesurfer-derived subcortical volume, cortical thickness, surface area, and local gyrification index in the PGC and DNS samples, respectively. T-PRS did not predict depression diagnosis (OR=1.007, 95%CI=[0.997-1.018]); however, it correlated with symptom severity in men (rho=0.175, p=7.957×10^−4^) in one large PGC cohort (N=762, 48% men). In DNS, T-PRS was associated with smaller amygdala volume in women (β=-0.186, t=-3.478, p=.001) and less prefrontal gyrification (max≤-2.970, p≤.006) in both sexes. In men, prefrontal gyrification mediated an indirect effect of T-PRS on broad depression risk (b=.005, p=.029), indexed using self-reported family history of depression. Depression-like shifts in cortical gene expression predict symptom severity in men and may contribute to disease vulnerability through their effect on cortical gyrification.

## INTRODUCTION

Major depressive disorder (MDD or ‘depression’) is a common and debilitating psychiatric illness characterized by low mood and anhedonia. Lifetime prevalence of MDD is estimated at up to 17% (1), and it constitutes the leading cause of disability worldwide (2). Despite the monumental impact of MDD, its biological bases remain incompletely understood.

Post-mortem gene expression studies have begun to shed light on the molecular mechanisms that may contribute to the emergence and maintenance of MDD. Most notably, a recent meta-analysis (3) of 8 post-mortem transcriptome datasets, including 51 depression cases and 50 matched controls, identified consistently altered expression of 566 ‘metaA-MDD’ genes across a corticolimbic circuit comprising the dorsolateral prefrontal cortex (DLPFC), subgenual anterior cingulate cortex (sgACC), and amygdala. Of these genes, 56% were downregulated in depression cases, relative to controls. Subsequent pathway analysis revealed associations with biological functions related to cell death and survival as well as cell-to-cell signaling, and suggested reduced neurotrophic support and GABA function.

Although these post-mortem findings provide important information about the molecular pathways whose dysregulation may underlie MDD pathophysiology, they offer little insight into how these microscale processes may impact larger-scale neural structure and function to ultimately contribute to symptom emergence. To address this important gap, our group developed a novel transcriptome-based polygenic risk score (T-PRS) that makes use of common genetic variants to translate these post-mortem molecular changes into an in vivo peripheral proxy measure capturing similarity to the post-mortem depression transcriptome. We recently mapped this score onto corticolimbic function in a sample of healthy young adults, associating higher T-PRS with a female-specific pattern of blunted reactivity to neutral faces further predictive of subclinical anhedonia (4). These findings suggest that molecular shifts towards a depression-like transcriptome may contribute to a functional risk phenotype even in the absence of clinically-significant depression. However, the impact of these molecular shifts on clinical MDD risk or more stable trait-like measures of brain morphology remains unknown.

To answer these important questions, we first sought to evaluate the association of our T-PRS to MDD diagnosis and symptom severity in a large and well-characterized sample obtained from the Psychiatric Genomics Consortium MDD working group (PGC-MDD) (5, 6). Second, to provide mechanistic insight into the biological pathways that may contribute to any association with clinical risk, we further sought to identify the effects of T-PRS on measures of brain morphology in a sample of healthy young adults participating in the Duke Neurogenetics Study. Finally, in order to identify convergence between analyses, we sought to link T-PRS-associated brain morphology phenotypes with self-reported family history of depression as a proxy for broad depression risk in our non-clinical sample.

To obtain a thorough characterization of the neuroanatomical signature of the T-PRS, we examined the effects of the PRS on volume in subcortical regions, as well as on cortical thickness and surface area. We also examined its effects on local gyrification, a less-studied cortical phenotype that develops in early life and has been linked to cortical complexity, across the cortex. By examining each of these brain-based phenotypes, which have unique genetic origins (7) and developmental and aging-related trajectories (8), we sought to capture links between depression-associated changes in gene expression and distinct variations in brain structure that could indicate atypical neurodevelopment (i.e., variations in cortical surface area or local gyrification) or accelerated aging (i.e., variations in cortical thickness or subcortical volume), both of which have been implicated in the pathophysiology of MDD (9, 10). Given consistent evidence of sex differences in depression risk (11, 12) and sex-specificity of T-PRS effects in our prior work (4), we tested associations between T-PRS and brain morphology in men and women separately.

Although we aimed to maintain a discovery component to this study, we held several predictions when testing our main hypotheses that T-PRS would be associated with MDD diagnosis and symptom severity and brain morphology. Above all, we expected to observe associations between T-PRS and morphology in corticolimbic regions, including those from which T-PRS was originally derived (3). Second, we expected to observe particularly strong negative associations between T-PRS and cortical surface area, a highly heritable morphological phenotype (7) that has been genetically linked to depression (13, 14).

## MATERIALS AND METHODS

### PGC Analyses

To validate the link between T-PRS and depression in a clinical sample, we computed T-PRS in 29,340 individuals (59% women; 12,923 MDD cases and 16,417 controls) from 21 cohorts included in the PGC. Only individuals with European genetic background were included in the sample, and principal components were derived for each individual and used to account for residual population substructure in each cohort. Calculation of the T-PRS was performed separately for each cohort and is described in depth below.

Logistic regressions, including sex and the aforementioned principal components accounting for residual population substructure as covariates, were used to test associations between T-PRS and depression outcome (case/control) in each of the 21 PGC cohorts. Similar regressions were fitted for each cohort, stratified by sex. One-sided p-values were obtained for each regression. Results from cohort-specific logistic regressions were used to perform a generic inverse variance meta-analysis using the META package in R. Three meta-analyses were performed: 1) not stratified by sex and including sex as a covariate; 2) stratified by sex (women only); 3) stratified by sex (men only).

Spearman correlation was used to test associations between T-PRS and depression symptom severity, assessed with the 21-item version of the Hamilton Depression Rating Scale (HDRS), in the subset of MDD cases for whom this data was available (n=762, 48% male, from the MARS cohort).

All analyses were performed in R.

### Neuroimaging sample

This study used archival data from 482 university students (226 men, 256 women; aged 19.78 ± 1.23 years) who participated in the Duke Neurogenetics Study (DNS). All participants provided informed consent in accordance with Duke University guidelines, and all were in good general health (for full exclusionary criteria, see (15). Participants were screened for DSM-IV Axis I disorders plus select DSM-IV Axis II disorders (Antisocial Personality Disorder, Borderline Personality Disorder) using the eMINI (16), but a current or lifetime diagnosis was not necessarily exclusionary. A complete list of participant diagnoses can be found in Supplementary Table 1. The Duke University Institutional Review Board approved all study procedures.

We restricted our analyses to non-Hispanic white participants to match the ethnic background of the post-mortem cohorts used to develop our T-PRS. Our sample was reduced to 478 subjects after assessing the presence of relatedness and population stratification, as described in the Supplemental Materials. Where appropriate, the first principal component (C1) from a multidimensional scaling (MDS) analysis (17), which accounted for 50% of variance, was included as a covariate to account for residual population substructure (20).

### Calculation of the transcriptome-based polygenic risk score

Methods for DNA extraction and genotyping were described in (21). Our T-PRS, which captures depression-related changes in gene expression, was developed based on a list of 566 genes generated by a meta-analysis of case-control post-mortem brain transcriptome datasets (n=101 post-mortem subjects; 51 MDD, 50 controls) (3). Using PrediXcan (22) and GTEx ‘cortex’ tissue as a reference transcriptome, we successfully imputed relative cortical expression of 76 out of these 566 genes at the individual participant level. Imputation was based on SNPs genotyped in peripheral tissue of DNS participants. Individual SNP contributions were determined based on weighting in a tissue-specific prediction model (gtex_v7_Brain_Cortex_imputed_europeans_tw_0.5_signif.db). Expression levels were not imputed for the remaining genes, which did not have significant cis-eQTLs identified in the reference tissue, due to low power and/or lower expression heritability (22). Once imputed, expression values were weighted by direction of effect in the original post-mortem meta-analysis (3), and T-PRS was computed as the sum of the weighted expression values of the 76 imputed genes, with higher values indicating a more depression-like transcriptome. We have provided a list of the 76 genes included in our T-PRS in Supplementary Table 2.

### Acquisition and preprocessing of MRI data

Each participant was scanned using one of the two identical research-dedicated GE MR750 3T scanners at the Duke-UNC Brain Imaging and Analysis Center. This scanner is equipped with high-power high-duty-cycle 50-mT/m gradients at 200 T/m/s slew rate and an eight-channel head coil for parallel imaging at high bandwidth up to 1⍰MHz. High-resolution 3D T1-weighted structural images were obtained using a 3D Ax FSPGR BRAVO sequence with the following parameters: TE=3.22⍰ms, TR=8.148⍰ms, FOV=240⍰mm, flip angle=12°, 162 sagittal slices, matrix=256×256, slice thickness=1⍰mm with no gap.

MRI data was processed using Freesurfer (http://surfer.nmr.mgh.harvard.edu, version 6.0). Pipelines and parameters are described in detail in Supplemental Materials.

### Volume- and surface-based analyses

Sex-specific main effects of T-PRS on subcortical volume were tested with separate linear regressions including age, estimated total intracranial volume (eTIV), and C1 as nuisance variables. To account for testing in multiple regions (n=7), a Bonferroni-adjusted threshold, p≤.0071 (.05/7), was used to determine significance.

Sex-specific main effects of T-PRS on vertex-wise CT, CSA, and LGI were tested with separate linear regressions including age, eTIV (for CSA only), and C1 as nuisance variables. Regions of interest (ROIs) were identified using a modified cluster-size exclusion method for multiple comparisons correction whereby cluster-wise probability was estimated using a Monte Carlo simulation with a vertex-wise threshold, p<.05, and 10,000 repetitions (23). To account for testing of multiple cortical phenotypes (n=3), a Bonferroni-adjusted threshold, p≤.0167 (.05/3), was used to determine cluster-wise significance.

### Links to familial depression

Sex-specific linear regressions, also fitted in R, were used to test associations between self-reported family history of depression (‘familial depression’) and covariate-adjusted morphology in each of the T-PRS-associated regions in the DNS sample (n=4 in men, 10 in women). Family history of depression was assessed using the following item, to which participants responded ‘yes’ or ‘no’: “Has anyone in your family ever felt sad, blue, or depressed for most of the time for two weeks or more?” For this question, family was defined as ‘immediate, biological family only (biological mother, biological father, biological brothers or sisters).’ Participants were asked to indicate whether they were ‘not confident at all’, ‘reasonably confident’, or ‘very confident’ in their responses to this question and others (“How confident do you generally feel about the information you have just given about your family members?”). Significance was determined using a Bonferroni-adjusted threshold, p≤.0125 (.05/4, men) or p≤.005 (.05/10, women). A causal mediation analysis, performed in R with nonparametric bootstrapping, was used to test relationships among T-PRS, cluster-wise cortical gyrification, and familial depression.

## RESULTS

### T-PRS association with MDD diagnosis and symptom severity

Meta-analyses including results from all 21 PGC cohorts did not reveal links between T-PRS and diagnosis in the entire sample (OR=1.007, 95% C.I.=[0.997 - 1.018]) or in the sex-stratified subsamples (OR=1.009, 95% C.I.=[0.996 - 1.023] in women; OR=1.004, 95% C.I.=[0.987 - 1.022] in men) (Supplementary Tables 4-6). However, T-PRS was correlated with greater symptom severity in male cases (rho=0.175, p=7.957 × 10^−4^, n=363), but not female cases (rho=0.024, p=0.638, n=399), in the only cohort where the HDRS was available (Figure 1). In the same cohort, T-PRS was nominally associated with increased risk for MDD in men (MARS 1: OR=1.073, p=0.038) (Supplementary Table 3), but not women (OR=0.991, p=0.607).

**Figure 1.**
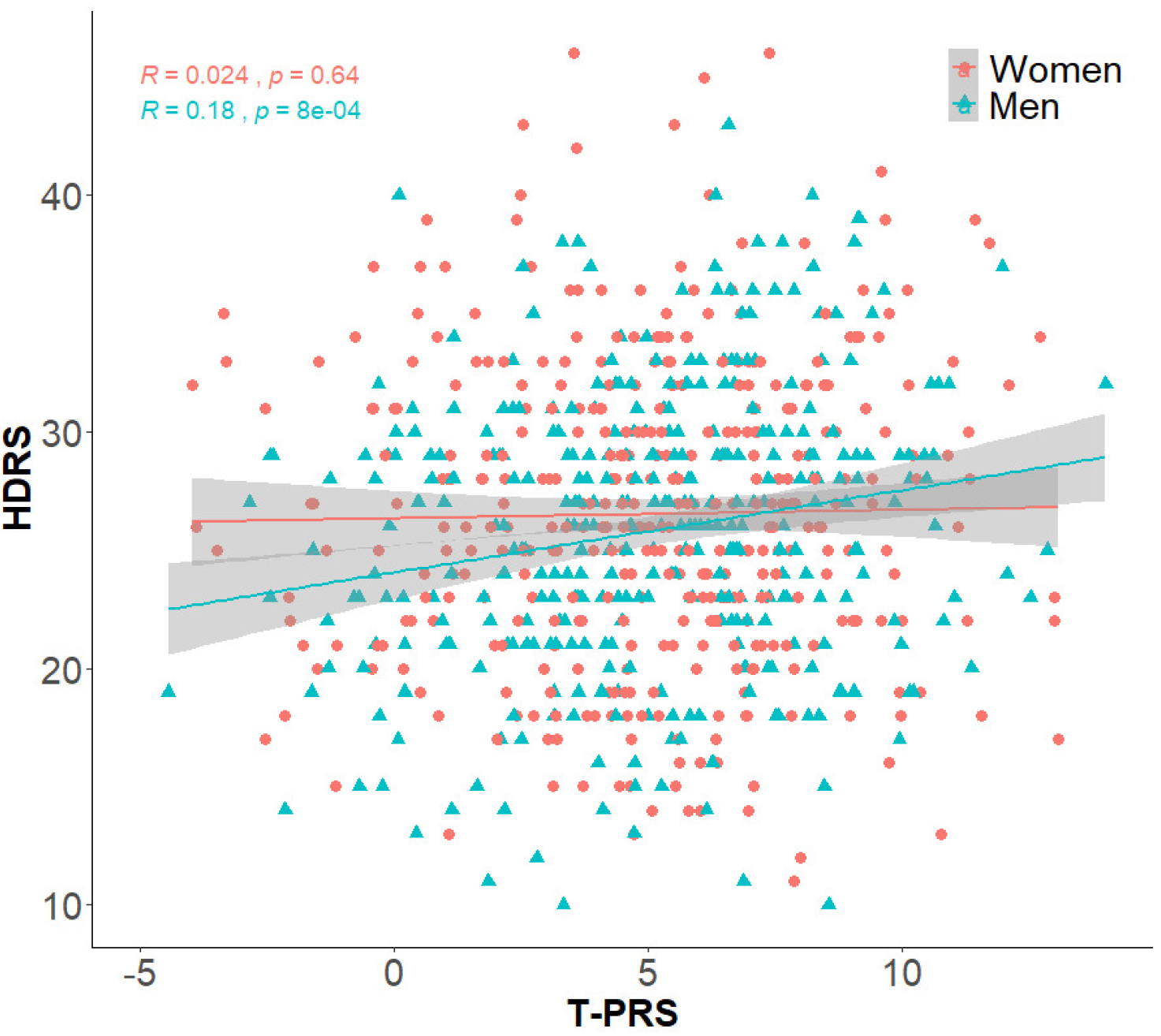
Correlation between T-PRS and depression symptoms in MARS cohorts. HDRS=Hamilton Depression Rating Scale

### Neuroanatomical signature of T-PRS

Effects of T-PRS on brain morphology also differed by sex. Specifically, in our neuroimaging analyses, T-PRS was associated with amygdala volume in women (Table 1) and a distinct patterns of vertex-wise local gyrification in both sexes (Supplementary Table 7, Figure 2). T-PRS was not a significant predictor of subcortical volume in men or of vertex-wise cortical thickness and cortical surface area in either sex.

**Table 1.** Sex-specific main effects of T-PRS on regional subcortical volume.

| Volume | Men |  | Women |  |
| --- | --- | --- | --- | --- |
|  | t | p (pcrit<.0071) | T | p (pcrit<.0071) |
| Thalamus | -1.783 | .076 | 1.450 | .148 |
| Caudate | -0.172 | .864 | -1.352 | .178 |
| Putamen | -0.888 | .375 | 2.215 | .028 |
| Pallidum | -0.409 | .683 | 0.164 | .869 |
| Hippocampus | -0.691 | .490 | -1.776 | .077 |
| Amygdala | 1.343 | .181 | **-3.478** | **.001** |
| Accumbens area | -2.294 | .023 | -0.739 | .461 |

*Bold indicates significance, as determined using pcrit<.0071*

**Figure 2.**
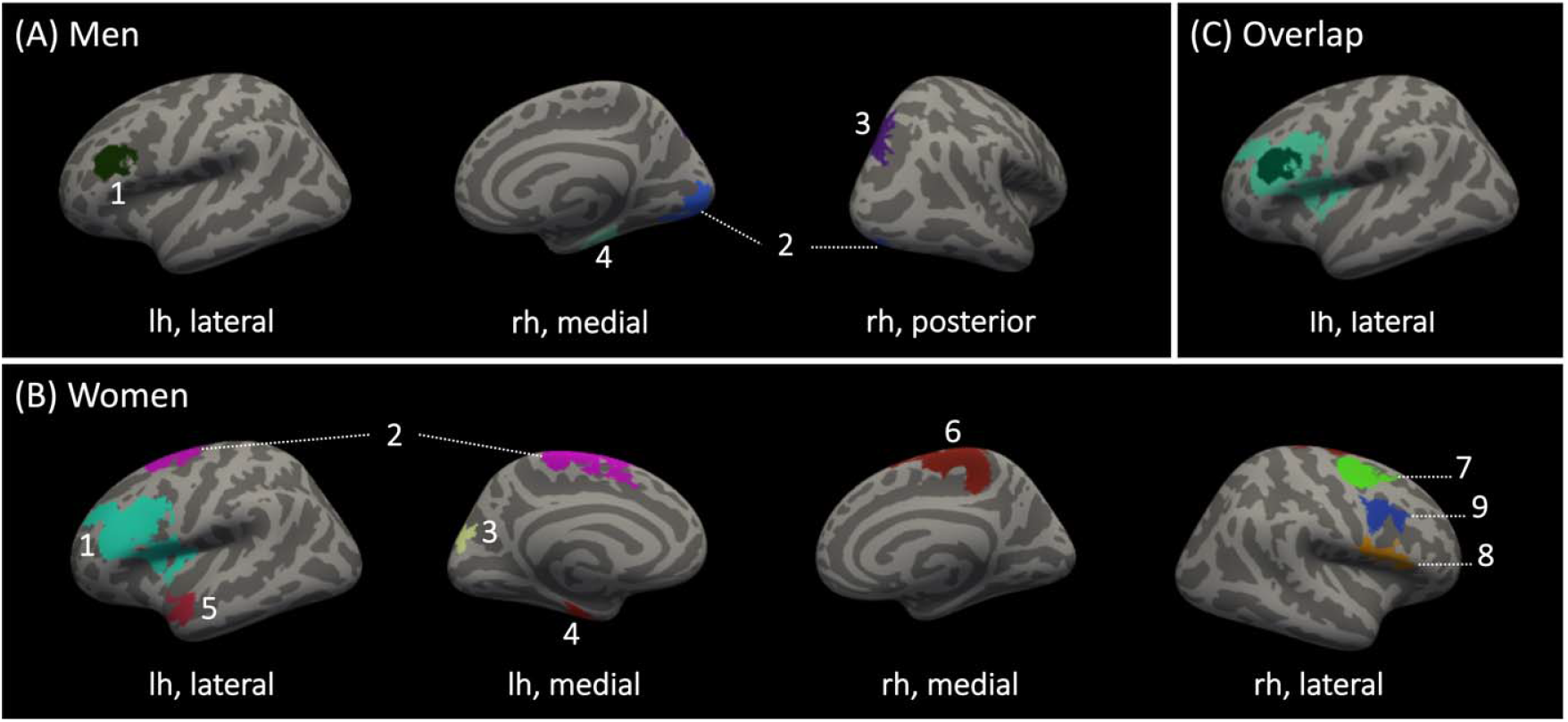
Clusters in which T-PRS was a significant predictor of vertex-wise local gyrification in men and women. LGI=local gyrification index; lh=left hemisphere; rh=right hemisphere (A)Clusters in which T-PRS was negatively associated (1,2) or positively associated (3,4) with LGI in men. (B) Clusters in which T-PRS was negatively associated with LGI in women. (C) Overlapping clusters in which T-PRS was negatively associated with LGI in men and women.

In women, higher T-PRS, indicative of a more depression-like brain transcriptome, was associated with less local gyrification (max≤-2.118, cluster-wise p≤.014) across large swaths of the cortex, including nine clusters with peak vertices in the bilateral prefrontal cortex (clusters #1, 2, 7, 8, 9), right paracentral cortex (cluster #6), left cuneus (cluster #3), left fusiform cortex (cluster #4), and left superior temporal cortex (cluster #5). Furthermore, higher T-PRS was also strongly associated with lower amygdala volume (β=-0.186, t=-3.478, p=.001). Each effect remained significant when tested with a follow-up linear regression including psychiatric diagnosis as an additional covariate (p≤.010, Supplementary Table 8) and when tested in a restricted sample, from which participants with a lifetime psychiatric diagnosis were excluded (n=198, p≤.011, Supplementary Table 8).

In men, higher T-PRS was associated with less local gyrification in two clusters (max≤-2.970, cluster-wise p≤.006), with peak vertices in the left rostral middle frontal cortex (cluster #1) and right lingual cortex (cluster #2). It was also associated with more local gyrification in two clusters (max≥2.109, cluster-wise p≤.008), with peak vertices in the right superior parietal cortex (cluster #3) and right fusiform cortex (cluster #4). In almost all cases, the effect of T-PRS on clusterwise gyrification remained significant when tested with a follow-up linear regression including psychiatric diagnosis as an additional covariate (p≤.024, Supplementary Table 8) and when tested in a restricted sample, from which participants with a lifetime psychiatric diagnosis were excluded (n=163, p≤.040 (excludes cluster #4), Supplementary Table 8).

There was substantial overlap between cluster #1 in men and cluster #1 in women. Gyrification in each of these clusters, which had peak vertices in the left rostral middle frontal cortex, was negatively associated with T-PRS. There was no additional overlap between T-PRS-associated clusters in men and women.

### Neuroanatomical link between T-PRS and familial depression

Familial depression was significantly associated with morphology in T-PRS-associated regions in men, but not in women (Supplementary Table 9). Specifically, in men, familial depression, like T-PRS, was negatively associated with gyrification in the left rostral middle frontal cortex (cluster #1: t=-2.566, p=.011). A causal mediation analysis further indicated that hypogyrification in this region mediates an indirect link between T-PRS and broader depression risk, indexed by self-reported family history of depression, in this otherwise healthy sample (Figure 3). This association remained significant (t=-2.509, p=.013) when retested in a restricted sample (n=218), from which participants were excluded if reporting low confidence in their response to the corresponding self-report item. Moreover, this cluster formed part of a broader region identified in a follow-up vertex-wise analysis, testing main effects of familial depression on LGI, independent of T-PRS (Figure 4). This analysis revealed widespread associations between familial depression and hypogyrification in men (max≤-2.354, cluster-wise p≤.028), but none in women.

**Figure 3.**
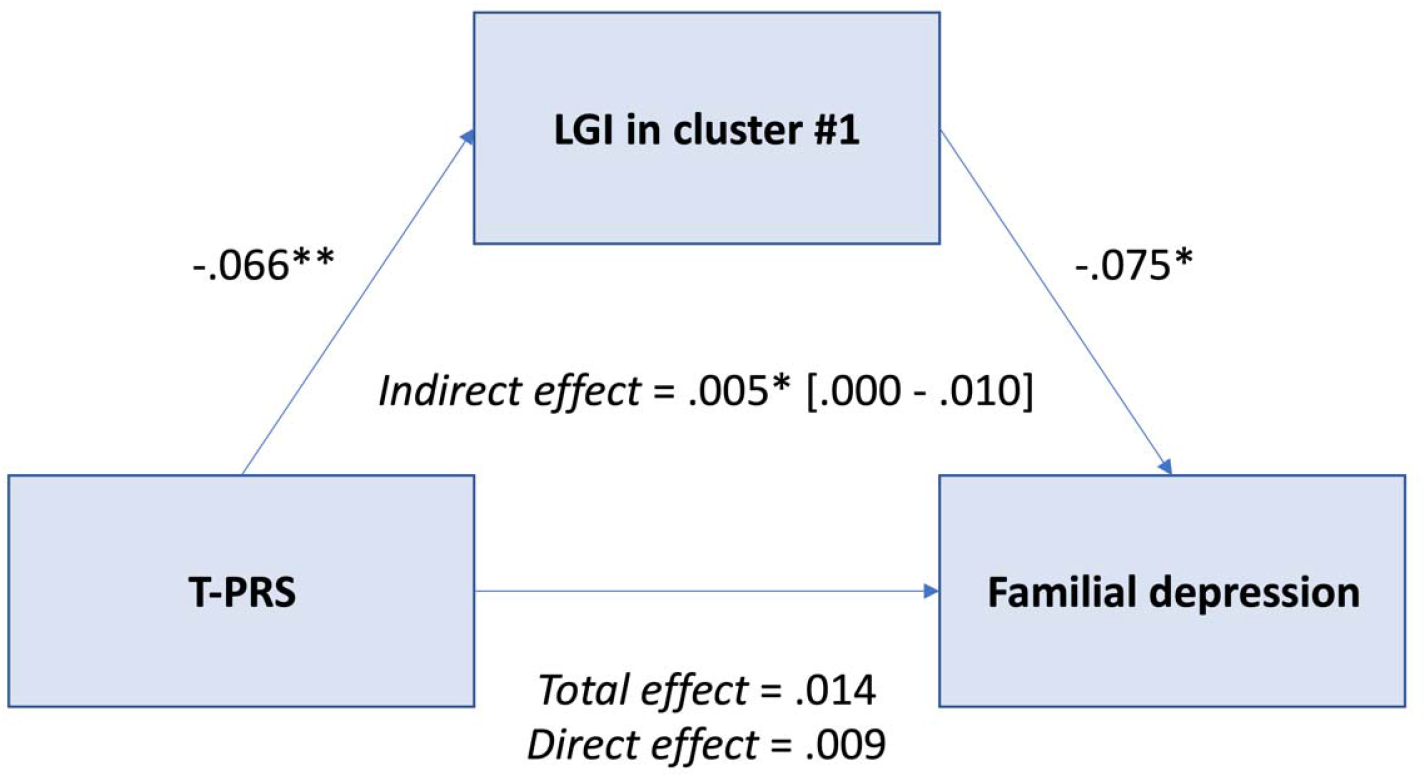
Indirect effect of T-PRS on broad depression risk, indexed by familial history of depression, in men. LGI=local gyrification index; p<.05 (*), p<01 (**) Unstandardized regression coefficients for the relationship between T-PRS and familial depression, as mediated by gyrification in the left rostral middle frontal cortex. The 95% confidence interval is in brackets.

**Figure 4.**
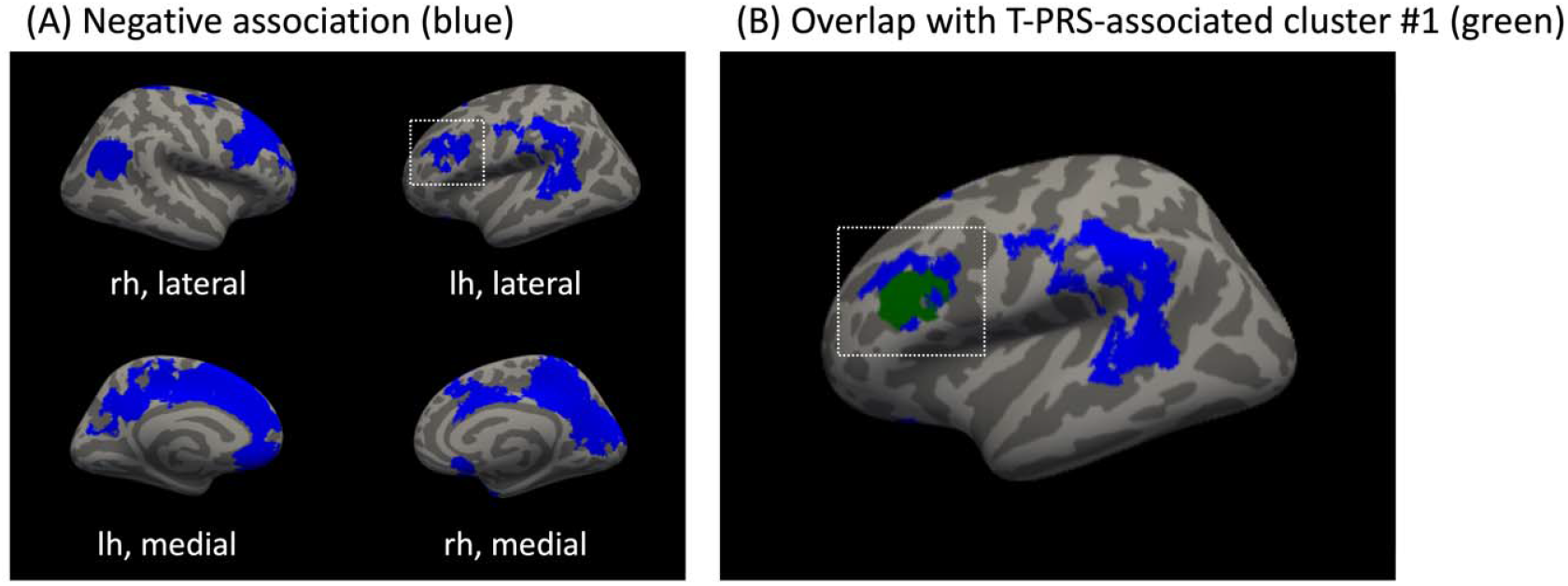
Clusters in which familial depression was a significant predictor of vertex-wise local gyrification in men. LGI=local gyrification index; lh=left hemisphere; rh=right hemisphere (A)Clusters in which familial depression was negatively associated with LGI in men: |max|=2.354 – 4.836, cluster-wise p≤.028. (B) Overlap with left rostral middle frontal cluster, in which T-PRS was negatively associated with LGI in men.

## DISCUSSION

In this study, we set out to identify the clinical and neuroanatomical correlates of a novel polygenic risk score (T-PRS) that captures common variants biasing gene expression towards a depression-like corticolimbic transcriptome. Although T-PRS was not a significant predictor of depression diagnosis in a large-scale clinical sample from the PGC-MDD, it was associated with clinical symptom severity in men with MDD. Moreover, we observed several significant relationships between T-PRS and discrete measures of brain morphology in a large non-clinical sample. First, we detected a significant negative association between T-PRS and amygdala volume in women. Second, we detected significant sex-specific associations between T-PRS and local gyrification. In women, higher T-PRS was associated with lower gyrification across large swaths of the bilateral frontal cortex. In men, higher T-PRS was associated with lower gyrification in lateral frontal and medial occipital regions and with higher gyrification in lateral parietal and medial temporal regions. Hypogyrification in the lateral frontal region in men was further associated with familial depression.

Our PGC analyses indicated that, among men with MDD, higher T-PRS was associated with greater depressive symptom severity. These results provide an important initial validation of our approach and suggest a potential clinical relevance of the T-PRS. Of note, although we previously showed partially convergent effects of T-PRS and PRS derived from MDD genome-wide association studies (GWAS) on brain function phenotypes, we also showed that the scores are uncorrelated (4). The genes included in our T-PRS are also distinct from those emerging from transcriptome-wide association studies (TWAS) of depression, which seek to map MDD-associated SNPs onto specific gene expression patterns (24). These findings suggest that T-PRS may reflect a pathway of depression risk that is at least partially distinct from pathways captured by conventional GWAS or TWAS approaches. This divergence may in turn account for the lack of a direct link between T-PRS and MDD diagnosis in the larger PGC sample. In line with the brain tissue transcriptomic origins of the T-PRS, our results suggest that this score may be more strongly associated with specific brain-based risk endophenotypes for MDD rather than the full syndrome, which may be more biologically heterogeneous.

Consistent with our preliminary hypotheses, neuroanatomical patterns associated with higher T-PRS show significant spatial overlap with corticolimbic regions, including the amygdala and DLPFC, from which T-PRS is derived. Moreover, these findings are at least partially consistent with previous imaging studies in depression. For example, although a range of depression-associated changes in amygdala volume have been reported, reduced amygdala volume has been reliably demonstrated in unmedicated patients, relative to healthy controls (25), and smaller amygdala volume has been linked to more depressive symptoms in young adults from a large-scale population-based study (26). Likewise, reduced local gyrification in middle frontal regions has been reported in depressed patients, relative to healthy controls, and reduced local gyrification in prefrontal regions has been linked to number of depressive episodes (27), which in turn has been linked to greater heritability (28). The latter finding is particularly noteworthy given that links between more depression-like cortical gene expression and reduced prefrontal gyrification, including in regions overlapping the DLPFC, were observed in both men and women.

Our findings are also partially consistent with recent meta-analyses from the ENIGMA-MDD consortium. In these two large-scale studies, Schmaal et al. identified several neuroanatomical variants, including reductions in hippocampal volume (29) and age-dependent reductions in frontotemporal cortical thickness (14) and frontal, visual, somatosensory, and somatomotor cortical surface area (14), that robustly discriminated between depressed patients and healthy controls. These differences were driven largely by patients with recurrent depression and/or earlier age of onset, both of which have been linked to greater heritability (28), suggesting that observed variations therein may form part of a vulnerability profile that precedes the onset of MDD. Evidence of a genetic correlation between MDD and cortical surface area, reported in a recent GWAS by the ENIGMA consortium (13), and of a longitudinal association between baseline cortical surface area and future depression relapse (30) provides additional support for the existence of a vulnerability phenotype characterized by variations in cortical surface architecture. This vulnerability profile could also include variations in local gyrification (31), similar to the ones reported in independent case-control studies (27, 32, 33, 34, 35, 36). Given that cortical surface area increases rapidly during the first two years of life (37) and is phenotypically correlated with gyrification (8), it is possible that one might detect similar associations with T-PRS, reminiscent of those observed by Schmaal et al. (14), in a larger sample. It is also possible that the specific genetic variants captured by T-PRS exert a unique or particularly meaningful influence on cortical folding patterns while the primarily non-overlapping genetic variants identified in the ENIGMA GWAS shape degree of cortical expansion.

Gyrification develops in early life, being nearly complete by the age of two, when differential rates of tissue growth give rise to cortical folding, thereby increasing the cortical surface area, and, by extension, the number of neurons in a limited cranial volume (38, 39). This patterning is thought to optimize connectivity between adjacent regions (40), and remains relatively stable into adulthood (8). Thus, cortical gyrification can serve as an index of early brain development, and it can convey information about disruptions to neurodevelopmental processes, effects of which may persist into adulthood. The phenotype specificity of our findings suggests a uniquely impactful role of depression-like cortical gene expression in early life, when associated genetic and non-genetic risk factors could contribute to alterations or disruptions in cortical expansion and subsequent cortical folding, thereby giving rise to the observed variations in site-specific local gyrification. Reduced neurotrophic support and/or alterations in biological functions related to cell death and survival and cell-to-cell signaling, each of which was associated with genes in T-PRS (3), could contribute to atypical neurodevelopment during this critical period and could partially explain the link between depression-like cortical gene expression and associated variations in gyrification. In turn, these variations in cortical folding could contribute to structural dysconnectivity, subtle but widespread evidence of which has been reported in MDD (41). By extension, the site-specificity of our findings suggests a uniquely impactful role of depression-like cortical gene expression on structural connectivity within certain regions, namely the left DLPFC, stimulation of which has been shown to modulate resting-state functional connectivity within a meso-cortico-limbic network and has been the primary focus of neuromodulatory treatment of depression (42).

The sex-specificity of our findings is intriguing and warrants further consideration. The GTEx sample from which the PrediXcan reference transcriptome is derived is disproportionately male. As such, imputation accuracy could be better for men than women, accounting for the sex-specific link to depression symptom severity in the PGC sample. However, this explanation does not account for the much more widespread associations between T-PRS and local gyrification in women than in men. It is possible that women are more susceptible to morphological variations related to changes in cortical gene expression due to their unique hormonal milieu or developmental trajectory. This hypothesis is consistent with evidence of sex-dependent effects of maternal perinatal stress exposure on offspring cortical gyrification in adulthood that was recently reported by our group (43) and provides additional support for sex differences in responsivity to early life perturbations. It is also possible that the same hormonal conditions or developmental trajectories that contribute to sex differences in magnitude of effect buffer against negative consequences thereof in women. For example, altered patterns of cortical gene expression during early neurodevelopment could exert diffuse effects on cortical morphology in women, whose brains expand more rapidly during this critical period of development than do male brains (44). However, resilience to some of the proximal effects of prenatal and early postnatal stress, which has also been demonstrated in women (45), could mitigate the depressogenic effects of these structural variations.

This study has several limitations. First, we did not observe a link between T-PRS and MDD diagnosis in the full PGC-MDD sample, but rather only detected a male-specific association with symptom severity. It is important to recall, however, that the T-PRS was developed as a translational tool to assess the impact of depression-associated post-mortem brain transcriptomic changes on in vivo brain structure and function, not to capture broad genetic contributions to MDD, meaning its primary aim was achieved nonetheless. Second, we were only able to reliably impute cortical gene expression of a subset of the original metaA-MDD genes. This was due primarily to poor convergence between SNPs included in the prediction model and SNPs captured by the genotyping platform used to analyse DNS dataset. However, it also reflects a limitation of PrediXcan, which imputes expression levels based on observed genetic variation in the reference dataset and is therefore not suitable for genes whose expression is not highly genetically-regulated. Despite this limitation, however, PrediXcan uses a robust machine learning approach to impute expression based on multiple SNPs, hence we have high confidence in the genes we were able to impute. Finally, we restricted our analyses to non-Hispanic white participants to match the demographic characteristics of the GTEx reference dataset and Ding et al. (3) post-mortem cohorts. While this decision likely increased our ability to detect statistically significant effects, it also limits the generalizability of our findings to other ethnic groups. Future work should incorporate larger multi-ethnic samples, including MDD cases and controls covering the full range of symptom severity, in order to improve generalizability across ethnicities and facilitate further assessment of clinical relevance.

Despite these limitations, our results support the translational and clinical validity of a novel transcriptome-based polygenic risk score (T-PRS) indexing “depression-like” cortical gene expression changes previously only accessible via post-mortem tissue analysis. We provide strong evidence of sex-specific effects of T-PRS on volume of the amygdala, a central corticolimbic node, and frontal cortical gyrification, variations in which could indicate atypical neurodevelopment and contribute to depression-associated connectivity deficits. Given the discovery component of this study, replication is critical, as is further examination of the aforementioned sex-differences and their possible developmental and clinical implications. To that end, future work should explore the extent to which effects of T-PRS on brain structure and MDD risk may be developmentally mediated and therefore possibly amenable to early intervention and prevention efforts crucial for reducing disease burden.

## Data Availability

Data are available upon reasonable request from the principal investigators.

## AUTHOR CONTRIBUTIONS

AEM contributed to study design, conducted all neuroimaging analyses, and wrote the manuscript with guidance from YSN. FCD performed the genetic data processing, conducted the analyses on PGC data, and contributed to manuscript editing. ES contributed to study design and manuscript editing. EMB; MER, AMM, MJA, GP, EC, MP, BTB, KOS, CML, LAJ, IJ, RU, JWS, RHP, DFL, JBP, MMW, JS, GL, BWJHP, DIB, SPH contributed with data to the PGC-MDD working group and revising the final version of the manuscript. The PGC-MDD working group provided data access and analytic support for the case-control analyses. ARH designed and conducted the parent protocol of the Duke Neurogenetics Study. YSN developed the study concept, oversaw all analyses and all stages of manuscript preparation.

