## Supplementary Information for "Novel polygenic risk score links depression-related cortical transcriptomic changes to brain morphology and symptom severity"

Naomi R Wray* 1, 2

Stephan Ripke* 3, 4, 5

Manuel Mattheisen* 6, 7, 8

Maciej Trzaskowski 1

Enda M Byrne 1

Abdel Abdellaoui 9

Mark J Adams 10

Esben Agerbo 11, 12, 13

Tracy M Air 14

Till F M Andlauer 15, 16

Silviu-Alin Bacanu 17

Marie Bækvad-Hansen 13, 18

Aartjan T F Beekman 19

Tim B Bigdeli 17, 20

Elisabeth B Binder 15, 21

Julien Bryois 22

Henriette N Buttenschøn 13, 23, 24

Jonas Bybjerg-Grauholm 13, 18

Na Cai 25, 26

Enrique Castelao 27

Jane Hvarregaard Christensen 8, 13, 24

Toni-Kim Clarke 10

Jonathan R I Coleman 28

Lucía Colodro-Conde 29

Baptiste Couvy-Duchesne 2, 30

Nick Craddock 31

Gregory E Crawford 32, 33

Gail Davies 34

Franziska Degenhardt 35

Eske M Derks 29

Nese Direk 36, 37

Conor V Dolan 9

Erin C Dunn 38, 39, 40

Thalia C Eley 28

Valentina Escott-Price 41

Farnush Farhadi Hassan Kiadeh 42

Hilary K Finucane 43, 44

Jerome C Foo 45

Andreas J Forstner 35, 46, 47, 48

Josef Frank 45

Héléna A Gaspar 28

Michael Gill 49

Fernando S Goes 50

Scott D Gordon 29

Jakob Grove 8, 13, 24, 51

Lynsey S Hall 10, 52

Christine Søholm Hansen 13, 18

Thomas F Hansen 53, 54, 55

Stefan Herms 35, 47

Ian B Hickie 56

Per Hoffmann 35, 47

Georg Homuth 57

Carsten Horn 58

Jouke-Jan Hottenga 9

David M Hougaard 13, 18

David M Howard 10, 28

Marcus Ising 59

Rick Jansen 19

Ian Jones 60

Lisa A Jones 61

Eric Jorgenson 62

James A Knowles 63

Isaac S Kohane 64, 65, 66

Julia Kraft 4

Warren W. Kretzschmar 67

Zoltán Kutalik 68, 69

Yihan Li 67

Penelope A Lind 29

Donald J MacIntyre 70, 71

Dean F MacKinnon 50

Robert M Maier 2

Wolfgang Maier 72

Jonathan Marchini 73

Hamdi Mbarek 9

Patrick McGrath 74

Peter McGuffin 28

Sarah E Medland 29

Divya Mehta 2, 75

Christel M Middeldorp 9, 76, 77

Evelin Mihailov 78

Yuri Milaneschi 19

Lili Milani 78

Francis M Mondimore 50

Grant W Montgomery 1

Sara Mostafavi 79, 80

Niamh Mullins 28

Matthias Nauck 81, 82

Bernard Ng 80

Michel G Nivard 9

Dale R Nyholt 83

Paul F O'Reilly 28

Hogni Oskarsson 84

Michael J Owen 60

Jodie N Painter 29

Carsten Bøcker Pedersen 11, 12, 13

Marianne Giørtz Pedersen 11, 12, 13

Roseann E Peterson 17, 85

Wouter J Peyrot 19

Giorgio Pistis 27

Danielle Posthuma 86, 87

Jorge A Quiroz 88

Per Qvist 8, 13, 24

John P Rice 89

Brien P. Riley 17

Margarita Rivera 28, 90

Saira Saeed Mirza 36

Robert Schoevers 91

Eva C Schulte 92, 93

Ling Shen 62

Jianxin Shi 94

Stanley I Shyn 95

Engilbert Sigurdsson 96

Grant C B Sinnamon 97

Johannes H Smit 19

Daniel J Smith 98

Hreinn Stefansson 99

Stacy Steinberg 99

Fabian Streit 45

Jana Strohmaier 45

Katherine E Tansey 100

Henning Teismann 101

Alexander Teumer 102

Wesley Thompson 13, 54, 103, 104

Pippa A Thomson 105

Thorgeir E Thorgeirsson 99

Matthew Traylor 106

Jens Treutlein 45

Vassily Trubetskoy 4

André G Uitterlinden 107

Daniel Umbricht 108

Sandra Van der Auwera 109

Albert M van Hemert 110

Alexander Viktorin 22

Peter M Visscher 1, 2

Yunpeng Wang 13, 54, 104

Bradley T. Webb 111

Shantel Marie Weinsheimer 13, 54

Jürgen Wellmann 101

Gonneke Willemsen 9

Stephanie H Witt 45

Yang Wu 1

Hualin S Xi 112

Jian Yang 2, 113

Futao Zhang 1

Volker Arolt 114

Bernhard T Baune 114, 115, 116

Klaus Berger 101

Dorret I Boomsma 9

Sven Cichon 35, 47, 117, 118

Udo Dannlowski 114

EJC de Geus 9, 119

J Raymond DePaulo 50

Enrico Domenici 120

Katharina Domschke 121, 122

Tõnu Esko 5, 78

Hans J Grabe 109

Steven P Hamilton 123

Caroline Hayward 124

Andrew C Heath 89

Kenneth S Kendler 17

Stefan Kloiber 59, 125, 126

Glyn Lewis 127

Qingqin S Li 128

Susanne Lucae 59

Pamela AF Madden 89

Patrik K Magnusson 22

Nicholas G Martin 29

Andrew M McIntosh 10, 34

Andres Metspalu 78, 129

Ole Mors 13, 130

Preben Bo Mortensen 11, 12, 13, 24

Bertram Müller-Myhsok 15, 131, 132

Merete Nordentoft 13, 133

Markus M Nöthen 35

Michael C O'Donovan 60

Sara A Paciga 134

Nancy L Pedersen 22

Brenda WJH Penninx 19

Roy H Perlis 38, 135

David J Porteous 105

James B Potash 136

Martin Preisig 27

Marcella Rietschel 45

Catherine Schaefer 62

Thomas G Schulze 45, 93, 137, 138, 139

Jordan W Smoller 38, 39, 40

Kari Stefansson 99, 140

Henning Tiemeier 36, 141, 142

Rudolf Uher 143

Henry Völzke 102

Myrna M Weissman 74, 144

Thomas Werge 13, 54, 145

Cathryn M Lewis* 28, 146

Douglas F Levinson* 147

Gerome Breen* 28, 148

Anders D Børglum* 8, 13, 24

Patrick F Sullivan* 22, 149, 1

1, Institute for Molecular Bioscience, The University of Queensland, Brisbane, QLD, AU

2, Queensland Brain Institute, The University of Queensland, Brisbane, QLD, AU

3, Analytic and Translational Genetics Unit, Massachusetts General Hospital, Boston, MA, US

4, Department of Psychiatry and Psychotherapy, Universitätsmedizin Berlin Campus Charité Mitte, Berlin, DE

5, Medical and Population Genetics, Broad Institute, Cambridge, MA, US

6, Department of Psychiatry, Psychosomatics and Psychotherapy, University of Wurzburg, Wurzburg, DE

7, Centre for Psychiatry Research, Department of Clinical Neuroscience, Karolinska Institutet, Stockholm, SE

8, Department of Biomedicine, Aarhus University, Aarhus, DK

9, Dept of Biological Psychology & EMGO+ Institute for Health and Care Research, Vrije Universiteit Amsterdam, Amsterdam, NL

10, Division of Psychiatry, University of Edinburgh, Edinburgh, GB

11, Centre for Integrated Register-based Research, Aarhus University, Aarhus, DK

12, National Centre for Register-Based Research, Aarhus University, Aarhus, DK

13, iPSYCH, The Lundbeck Foundation Initiative for Integrative Psychiatric Research,, DK

14, Discipline of Psychiatry, University of Adelaide, Adelaide, SA, AU

15, Department of Translational Research in Psychiatry, Max Planck Institute of Psychiatry, Munich, DE

16, Department of Neurology, Klinikum rechts der Isar, Technical University of Munich, Munich, DE

17, Department of Psychiatry, Virginia Commonwealth University, Richmond, VA, US

18, Center for Neonatal Screening, Department for Congenital Disorders, Statens Serum Institut, Copenhagen, DK

19, Department of Psychiatry, Vrije Universiteit Medical Center and GGZ inGeest, Amsterdam, NL

20, Virginia Institute for Psychiatric and Behavior Genetics, Richmond, VA, US

21, Department of Psychiatry and Behavioral Sciences, Emory University School of Medicine, Atlanta, GA, US

22, Department of Medical Epidemiology and Biostatistics, Karolinska Institutet, Stockholm, SE

23, Department of Clinical Medicine, Translational Neuropsychiatry Unit, Aarhus University, Aarhus, DK

24, iSEQ, Centre for Integrative Sequencing, Aarhus University, Aarhus, DK

25, Human Genetics, Wellcome Trust Sanger Institute, Cambridge, GB

26, Statistical genomics and systems genetics, European Bioinformatics Institute (EMBL-EBI), Cambridge, GB

27, Department of Psychiatry, Lausanne University Hospital and University of Lausanne, Lausanne, CH

28, Social, Genetic and Developmental Psychiatry Centre, King's College London, London, GB

29, Genetics and Computational Biology, QIMR Berghofer Medical Research Institute, Brisbane, QLD, AU

30, Centre for Advanced Imaging, The University of Queensland, Brisbane, QLD, AU

31, Psychological Medicine, Cardiff University, Cardiff, GB

32, Center for Genomic and Computational Biology, Duke University, Durham, NC, US

33, Department of Pediatrics, Division of Medical Genetics, Duke University, Durham, NC, US

34, Centre for Cognitive Ageing and Cognitive Epidemiology, University of Edinburgh, Edinburgh, GB

35, Institute of Human Genetics, University of Bonn, School of Medicine & University Hospital Bonn, Bonn, DE

36, Epidemiology, Erasmus MC, Rotterdam, Zuid-Holland, NL

37, Psychiatry, Dokuz Eylul University School Of Medicine, Izmir, TR

38, Department of Psychiatry, Massachusetts General Hospital, Boston, MA, US

39, Psychiatric and Neurodevelopmental Genetics Unit (PNGU), Massachusetts General Hospital, Boston, MA, US

40, Stanley Center for Psychiatric Research, Broad Institute, Cambridge, MA, US

41, Neuroscience and Mental Health, Cardiff University, Cardiff, GB

42, Bioinformatics, University of British Columbia, Vancouver, BC, CA

43, Department of Epidemiology, Harvard T.H. Chan School of Public Health, Boston, MA, US

44, Department of Mathematics, Massachusetts Institute of Technology, Cambridge, MA, US

45, Department of Genetic Epidemiology in Psychiatry, Central Institute of Mental Health,  Medical Faculty Mannheim, Heidelberg University, Mannheim, Baden-Württemberg, DE

46, Department of Psychiatry (UPK), University of Basel, Basel, CH

47, Department of Biomedicine, University of Basel, Basel, CH

48, Centre for Human Genetics, University of Marburg, Marburg, DE

49, Department of Psychiatry, Trinity College Dublin, Dublin, IE

50, Psychiatry & Behavioral Sciences, Johns Hopkins University, Baltimore, MD, US

51, Bioinformatics Research Centre, Aarhus University, Aarhus, DK

52, Institute of Genetic Medicine, Newcastle University, Newcastle upon Tyne, GB

53, Danish Headache Centre, Department of Neurology, Rigshospitalet, Glostrup, DK

54, Institute of Biological Psychiatry, Mental Health Center Sct. Hans, Mental Health Services Capital Region of Denmark, Copenhagen, DK

55, iPSYCH, The Lundbeck Foundation Initiative for Psychiatric Research, Copenhagen, DK

56, Brain and Mind Centre, University of Sydney, Sydney, NSW, AU

57, Interfaculty Institute for Genetics and Functional Genomics, Department of Functional Genomics, University Medicine and Ernst Moritz Arndt University Greifswald, Greifswald, Mecklenburg-Vorpommern, DE

58, Roche Pharmaceutical Research and Early Development, Pharmaceutical Sciences, Roche Innovation Center Basel, F. Hoffmann-La Roche Ltd, Basel, CH

59, Max Planck Institute of Psychiatry, Munich, DE

60, MRC Centre for Neuropsychiatric Genetics and Genomics, Cardiff University, Cardiff, GB

61, Department of Psychological Medicine, University of Worcester, Worcester, GB

62, Division of Research, Kaiser Permanente Northern California, Oakland, CA, US

63, Psychiatry & The Behavioral Sciences, University of Southern California, Los Angeles, CA, US

64, Department of Biomedical Informatics, Harvard Medical School, Boston, MA, US

65, Department of Medicine, Brigham and Women's Hospital, Boston, MA, US

66, Informatics Program, Boston Children's Hospital, Boston, MA, US

67, Wellcome Trust Centre for Human Genetics, University of Oxford, Oxford, GB

68, Institute of Social and Preventive Medicine (IUMSP), Lausanne University Hospital and University of Lausanne, Lausanne, VD, CH

69, Swiss Institute of Bioinformatics, Lausanne, VD, CH

70, Division of Psychiatry, Centre for Clinical Brain Sciences, University of Edinburgh, Edinburgh, GB

71, Mental Health, NHS 24, Glasgow, GB

72, Department of Psychiatry and Psychotherapy, University of Bonn, Bonn, DE

73, Statistics, University of Oxford, Oxford, GB

74, Psychiatry, Columbia University College of Physicians and Surgeons, New York, NY, US

75, School of Psychology and Counseling, Queensland University of Technology, Brisbane, QLD, AU

76, Child and Youth Mental Health Service, Children's Health Queensland Hospital and Health Service, South Brisbane, QLD, AU

77, Child Health Research Centre, University of Queensland, Brisbane, QLD, AU

78, Estonian Genome Center, University of Tartu, Tartu, EE

79, Medical Genetics, University of British Columbia, Vancouver, BC, CA

80, Statistics, University of British Columbia, Vancouver, BC, CA

81, DZHK (German Centre for Cardiovascular Research), Partner Site Greifswald, University Medicine, University Medicine Greifswald, Greifswald, Mecklenburg-Vorpommern, DE

82, Institute of Clinical Chemistry and Laboratory Medicine, University Medicine Greifswald, Greifswald, Mecklenburg-Vorpommern, DE

83, Institute of Health and Biomedical Innovation, Queensland University of Technology, Brisbane, QLD, AU

84, Humus, Reykjavik, IS

85, Virginia Institute for Psychiatric & Behavioral Genetics, Virginia Commonwealth University, Richmond, VA, US

86, Clinical Genetics, Vrije Universiteit Medical Center, Amsterdam, NL

87, Complex Trait Genetics, Vrije Universiteit Amsterdam, Amsterdam, NL

88, Solid Biosciences, Boston, MA, US

89, Department of Psychiatry, Washington University in Saint Louis School of Medicine, Saint Louis, MO, US

90, Department of Biochemistry and Molecular Biology II, Institute of Neurosciences, Biomedical Research Center (CIBM), University of Granada, Granada, ES

91, Department of Psychiatry, University of Groningen, University Medical Center Groningen, Groningen, NL

92, Department of Psychiatry and Psychotherapy, University Hospital, Ludwig Maximilian University Munich, Munich, DE

93, Institute of Psychiatric Phenomics and Genomics (IPPG), University Hospital, Ludwig Maximilian University Munich, Munich, DE

94, Division of Cancer Epidemiology and Genetics, National Cancer Institute, Bethesda, MD, US

95, Behavioral Health Services, Kaiser Permanente Washington, Seattle, WA, US

96, Faculty of Medicine, Department of Psychiatry, University of Iceland, Reykjavik, IS

97, School of Medicine and Dentistry, James Cook University, Townsville, QLD, AU

98, Institute of Health and Wellbeing, University of Glasgow, Glasgow, GB

99, deCODE Genetics / Amgen, Reykjavik, IS

100, College of Biomedical and Life Sciences, Cardiff University, Cardiff, GB

101, Institute of Epidemiology and Social Medicine, University of Münster, Münster, Nordrhein-Westfalen, DE

102, Institute for Community Medicine, University Medicine Greifswald, Greifswald, Mecklenburg-Vorpommern, DE

103, Department of Psychiatry, University of California, San Diego, San Diego, CA, US

104, KG Jebsen Centre for Psychosis Research, Norway Division of Mental Health and Addiction, Oslo University Hospital, Oslo, NO

105, Medical Genetics Section, CGEM, IGMM, University of Edinburgh, Edinburgh, GB

106, Clinical Neurosciences, University of Cambridge, Cambridge, GB

107, Internal Medicine, Erasmus MC, Rotterdam, Zuid-Holland, NL

108, Roche Pharmaceutical Research and Early Development, Neuroscience, Ophthalmology and Rare Diseases Discovery & Translational Medicine Area, Roche Innovation Center Basel, F. Hoffmann-La Roche Ltd, Basel, CH

109, Department of Psychiatry and Psychotherapy, University Medicine Greifswald, Greifswald, Mecklenburg-Vorpommern, DE

110, Department of Psychiatry, Leiden University Medical Center, Leiden, NL

111, Virginia Institute for Psychiatric & Behavioral Genetics, Virginia Commonwealth University, Richmond, VA, US

112, Computational Sciences Center of Emphasis, Pfizer Global Research and Development, Cambridge, MA, US

113, Institute for Molecular Bioscience; Queensland Brain Institute, The University of Queensland, Brisbane, QLD, AU

114, Department of Psychiatry, University of Münster, Münster, Nordrhein-Westfalen, DE

115, Department of Psychiatry, Melbourne Medical School, University of Melbourne, Melbourne, AU

116, Florey Institute for Neuroscience and Mental Health, University of Melbourne, Melbourne, AU

117, Institute of Medical Genetics and Pathology, University Hospital Basel, University of Basel, Basel, CH

118, Institute of Neuroscience and Medicine (INM-1), Research Center Juelich, Juelich, DE

119, Amsterdam Public Health Institute, Vrije Universiteit Medical Center, Amsterdam, NL

120, Centre for Integrative Biology, Università degli Studi di Trento, Trento, Trentino-Alto Adige, IT

121, Department of Psychiatry and Psychotherapy, Medical Center - University of Freiburg, Faculty of Medicine, University of Freiburg, Freiburg, DE

122, Center for NeuroModulation, Faculty of Medicine, University of Freiburg, Freiburg, DE

123, Psychiatry, Kaiser Permanente Northern California, San Francisco, CA, US

124, Medical Research Council Human Genetics Unit, Institute of Genetics and Molecular Medicine, University of Edinburgh, Edinburgh, GB

125, Department of Psychiatry, University of Toronto, Toronto, ON, CA

126, Centre for Addiction and Mental Health, Toronto, ON, CA

127, Division of Psychiatry, University College London, London, GB

128, Neuroscience Therapeutic Area, Janssen Research and Development, LLC, Titusville, NJ, US

129, Institute of Molecular and Cell Biology, University of Tartu, Tartu, EE

130, Psychosis Research Unit, Aarhus University Hospital, Risskov, Aarhus, DK

131, Munich Cluster for Systems Neurology (SyNergy), Munich, DE

132, University of Liverpool, Liverpool, GB

133, Mental Health Center Copenhagen, Copenhagen Universtity Hospital, Copenhagen, DK

134, Human Genetics and Computational Biomedicine, Pfizer Global Research and Development, Groton, CT, US

135, Psychiatry, Harvard Medical School, Boston, MA, US

136, Psychiatry, University of Iowa, Iowa City, IA, US

137, Department of Psychiatry and Behavioral Sciences, Johns Hopkins University, Baltimore, MD, US

138, Department of Psychiatry and Psychotherapy, University Medical Center Göttingen, Goettingen, Niedersachsen, DE

139, Human Genetics Branch, NIMH Division of Intramural Research Programs, Bethesda, MD, US

140, Faculty of Medicine, University of Iceland, Reykjavik, IS

141, Child and Adolescent Psychiatry, Erasmus MC, Rotterdam, Zuid-Holland, NL

142, Psychiatry, Erasmus MC, Rotterdam, Zuid-Holland, NL

143, Psychiatry, Dalhousie University, Halifax, NS, CA

144, Division of Translational Epidemiology, New York State Psychiatric Institute, New York, NY, US

145, Department of Clinical Medicine, University of Copenhagen, Copenhagen, DK

146, Department of Medical & Molecular Genetics, King's College London, London, GB

147, Psychiatry & Behavioral Sciences, Stanford University, Stanford, CA, US

148, NIHR Maudsley Biomedical Research Centre, King's College London, London, GB

149, Genetics, University of North Carolina at Chapel Hill, Chapel Hill, NC, US

150, Psychiatry, University of North Carolina at Chapel Hill, Chape

**SUPPLEMENTAL MATERIALS**

*Neuroimaging sample*

We restricted our analyses to non-Hispanic white participants to match the ethnic background of the postmortem cohorts used to develop our T-PRS, and we further probed the resulting population structure as follows. First, we used proportional identity by descent (PIHAT) to assess the presence of relatedness in the sample. When pairs of individuals with PIHAT > 0.2 were detected, we excluded from the analysis the individual with higher SNP-level missingness (i.e. lower genome-wide call rate). Using this method, we identified two sibling pairs (PIHAT = 0.47, 0.49), and we reduced our sample to 480 subjects. Second, we used a multidimensional scaling (MDS) and clustering method (17), implemented in PLINK v1.9 (18) and using thresholds and recommendations published elsewhere (19, 20), to assess population stratification that remained after filtering by self-reported ethnicity. Two individuals from our sample did not cluster with the 1000 Genomes participants of European descent, and they were excluded from subsequent analyses, thereby reducing our sample to 478 subjects. A second MDS analysis yielded 10 principal components, the first of which (C1) explained almost 50% of variance. Where appropriate, this component was included as a covariate to account for residual population substructure (20).

*MRI preprocessing*

The Freesurfer automated segmentation pipeline (http://surfer.nmr.mgh.harvard.edu, version 6.0) was used to estimate volume in each of seven subcortical regions: accumbens area, amygdala, caudate, hippocampus, pallidum, putamen, and thalamus. This method has been described in detail elsewhere (46, 47). In brief, it performs: registration to standard space, intensity inhomogeneity correction, removal of non-brain tissue, tissue-type classification, and probabilistic anatomical labeling. Regional volumes were averaged over hemisphere to reduce the number of comparisons.

The Freesurfer surface-based processing stream (http://surfer.nmr.mgh.harvard.edu, version 6.0) was used to estimate cortical thickness (CT), cortical surface area (CSA), and local gyrification, quantified using the local gyrification index (LGI) (31), at each vertex on the cortical surface. This method has been described in detail elsewhere (31, 48, 49). In brief, it involves: defining the boundaries between white matter, grey matter, and cerebrospinal fluid; performing spherical transformation and areal interpolation; measuring the distance between white and pial surfaces (CT); measuring surface area on the inflated sphere (CSA); and quantifying the proportion of concealed versus visible pial surface area (LGI).

Once computed, vertex-wise estimates of cortical surface architecture were registered to the Freesurfer average template and smoothed with a Gaussian kernel (FWHM = 15mm). The latter step was not applied to vertex-wise estimates of LGI as they had comparable intrinsic smoothness. To ensure accuracy, all segmented volumes and reconstructed surfaces were assessed with Freesurfer quality assurance tools and visually inspected by a trained examiner, A.M. LGI computation failed in six subjects, who were excluded from the final sample (n = 472: 221 men, 251 women; aged 19.78 ± 1.24 years).

stylefix

**Supplementary Table 1. Participants in the original sample (n=482) meeting criteria for at least one DSM-IV Axis I diagnosis**

| **DSM-IV Axis I diagnosis** | **n** |
| --- | --- |
| Agoraphobia (with or without history of Panic Disorder) | 12 |
| Alcohol Abuse | 35 |
| Alcohol Dependence | 30 |
| Bipolar Disorder (past) | 16 |
| Generalized Anxiety Disorder | 5 |
| Major Depressive Disorder (current or past) | 26 |
| Obsessive Compulsive Disorder | 6 |
| Social Anxiety Disorder | 5 |
| Substance Abuse (cannabis) | 13 |
| Substance Dependence (cannabis) | 7 |
| **Total** | **114** |

**Supplementary Table 2. List of genes included in our T-PRS (n=76)**

| ACOT8 | DPY19L1 | PPP3CC |
| --- | --- | --- |
| ADH5 | DTNBP1 | PRKRIP1 |
| AGA | EIF4G3 | PRSS3 |
| AGL | FAM149A | RABGEF1 |
| ALDH4A1 | FIGNL1 | RPA1 |
| ALMS1 | GALNT13 | RPS26 |
| ANKRD10 | GAS2L1 | RRM1 |
| ARMC1 | GCC2 | RWDD2B |
| ARSA | GGCX | SFI1 |
| ATF4 | GOSR1 | SFT2D1 |
| ATIC | GPR98 | SIN3B |
| ATPIF1 | HN1L | SLC1A1 |
| BPHL | IVNS1ABP | SNX24 |
| BRMS1 | KCNIP3 | SPATA7 |
| CAMK2N2 | KIAA1467 | SPHK2 |
| CCNY | KLHL24 | STARD10 |
| CHERP | LXN | SYT7 |
| CIAO1 | MAPK9 | TAF1C |
| CLCN3 | MMACHC | TBCD |
| COQ5 | NEK1 | TMEM86B |
| CPNE7 | NPHP3 | TRAF3 |
| CSRP2 | PIK3R1 | TTC3 |
| DDT | PILRB | WWP2 |
| DGCR2 | PPIC | ZNF558 |
| DIDO1 | PPM1D |  |
| DOK4 | PPP2R3A |  |

**Supplementary Table 3. Generalized linear model results by cohort for T-PRS and the interaction between T-PRS and sex**

[see next three pages]

*N = sample size; N case = sample size in MDD group; N control = sample size in control group; OR = odds ratio for T-PRS in the GLM; 95% C.I. = 95% confidence interval of T-PRS odds ratio in the GLM; P 1s = one-sided p-value of T-PRS in the GLM; (T-PRS*Sex) = interaction term for T-PRS and sex in the GLM*

| **Cohort** | **Cohort name** | **Sex** | **N** | **N case** | **N control** | **O.R.** | **95% C.I.** | **P 1s** | **O.R. (T-PRS*Sex)** | **95% C.I. (T-PRS*Sex)** | **P (T-PRS*Sex)** |
| --- | --- | --- | --- | --- | --- | --- | --- | --- | --- | --- | --- |
| **1** | **CoFaMS** | **All** | 247 | 120 | 127 | 0.997 | 0.886-1.122 | 0.518 | 0.973 | 0.764-1.237 | 0.823 |
|  |  | **Women** | 135 | 74 | 61 | 1.008 | 0.866-1.175 | 0.457 |  |  |  |
|  |  | **Men** | 112 | 46 | 66 | 0.981 | 0.812-1.182 | 0.58 |  |  |  |
| **2** | **PsyCoLaus** | **All** | 1953 | 508 | 1445 | 1.012 | 0.954-1.073 | 0.347 | 1.089 | 0.964-1.231 | 0.173 |
|  |  | **Women** | 977 | 360 | 617 | 0.984 | 0.914-1.058 | 0.67 |  |  |  |
|  |  | **Men** | 976 | 148 | 828 | 1.067 | 0.968-1.178 | 0.097 |  |  |  |
| **3** | **Edinburgh** | **All** | 658 | 372 | 286 | 1.031 | 0.963-1.105 | 0.192 | 0.985 | 0.857-1.132 | 0.836 |
|  |  | **Women** | 360 | 221 | 139 | 1.036 | 0.946-1.136 | 0.224 |  |  |  |
|  |  | **Men** | 298 | 151 | 147 | 1.025 | 0.922-1.14 | 0.326 |  |  |  |
| **4** | **GenRED2** | **All** | 1305 | 831 | 474 | 1.012 | 0.966-1.061 | 0.303 | 1.053 | 0.955-1.162 | 0.303 |
|  |  | **Women** | 919 | 687 | 232 | 0.996 | 0.941-1.055 | 0.553 |  |  |  |
|  |  | **Men** | 386 | 144 | 242 | 1.046 | 0.965-1.133 | 0.138 |  |  |  |
| **5** | **STAR*D** | **All** | 1866 | 936 | 930 | 0.984 | 0.933-1.038 | 0.722 | 0.976 | 0.876-1.087 | 0.657 |
|  |  | **Women** | 983 | 558 | 425 | 0.995 | 0.925-1.07 | 0.552 |  |  |  |
|  |  | **Men** | 883 | 378 | 505 | 0.971 | 0.897-1.051 | 0.765 |  |  |  |
| **6** | **GSK/MPIP** | **All** | 1784 | 922 | 862 | 1.011 | 0.972-1.052 | 0.29 | 0.992 | 0.912-1.08 | 0.86 |
|  |  | **Women** | 1198 | 614 | 584 | 1.014 | 0.966-1.064 | 0.287 |  |  |  |
|  |  | **Men** | 586 | 308 | 278 | 1.007 | 0.939-1.079 | 0.426 |  |  |  |
| **7** | **MARS 1** | **All** | 1092 | 556 | 536 | 1.027 | 0.976-1.08 | 0.154 | 1.089 | 0.983-1.207 | 0.104 |
|  |  | **Women** | 584 | 289 | 295 | 0.991 | 0.925-1.06 | 0.607 |  |  |  |
|  |  | **Men** | 508 | 267 | 241 | 1.073 | 0.993-1.16 | **0.038** |  |  |  |
| **8** | **MARS 2** | **All** | 602 | 251 | 351 | 1.017 | 0.945-1.094 | 0.33 | 1.045 | 0.901-1.213 | 0.565 |
|  |  | **Women** | 302 | 131 | 171 | 1.008 | 0.913-1.113 | 0.437 |  |  |  |
|  |  | **Men** | 300 | 120 | 180 | 1.048 | 0.936-1.177 | 0.211 |  |  |  |
| **9** | **QIMR 1** | **All** | 967 | 582 | 385 | 0.955 | 0.899-1.013 | 0.937 | 0.935 | 0.827-1.056 | 0.281 |
|  |  | **Women** | 566 | 328 | 238 | 0.98 | 0.908-1.057 | 0.703 |  |  |  |
|  |  | **Men** | 401 | 254 | 147 | 0.917 | 0.832-1.009 | 0.962 |  |  |  |
| **10** | **QIMR 2** | **All** | 1088 | 499 | 589 | 1.014 | 0.967-1.063 | 0.285 | 0.946 | 0.857-1.045 | 0.275 |
|  |  | **Women** | 712 | 347 | 365 | 1.034 | 0.975-1.098 | 0.134 |  |  |  |
|  |  | **Men** | 376 | 152 | 224 | 0.979 | 0.904-1.059 | 0.702 |  |  |  |
| **11** | **QIMR 3** | **All** | 1091 | 565 | 526 | 0.992 | 0.931-1.056 | 0.602 | 0.887 | 0.774-1.017 | 0.086 |
|  |  | **Women** | 707 | 404 | 303 | 1.031 | 0.954-1.114 | 0.22 |  |  |  |
|  |  | **Men** | 384 | 161 | 223 | 0.924 | 0.825-1.033 | 0.916 |  |  |  |
| **12** | **RADIANT-UK** | **All** | 3253 | 1872 | 1381 | 1.024 | 0.995-1.054 | 0.052 | 1.02 | 0.961-1.083 | 0.513 |
|  |  | **Women** | 2146 | 1323 | 823 | 1.017 | 0.981-1.055 | 0.173 |  |  |  |
|  |  | **Men** | 1107 | 549 | 558 | 1.036 | 0.988-1.087 | 0.073 |  |  |  |
| **13** | **RADIANT-GER** | **All** | 774 | 327 | 447 | 1.029 | 0.968-1.095 | 0.18 | 0.931 | 0.819-1.059 | 0.278 |
|  |  | **Women** | 437 | 215 | 222 | 1.054 | 0.975-1.14 | 0.093 |  |  |  |
|  |  | **Men** | 337 | 112 | 225 | 0.986 | 0.889-1.093 | 0.604 |  |  |  |
| **14** | **RADIANT-US** | **All** | 600 | 223 | 377 | 1.054 | 0.98-1.134 | 0.08 | 1.019 | 0.869-1.195 | 0.819 |
|  |  | **Women** | 371 | 175 | 196 | 1.048 | 0.961-1.145 | 0.146 |  |  |  |
|  |  | **Men** | 229 | 48 | 181 | 1.062 | 0.929-1.216 | 0.19 |  |  |  |
| **15** | **TwinGene** | **All** | 3760 | 1098 | 2662 | 1.007 | 0.979-1.035 | 0.316 | 0.952 | 0.899-1.009 | 0.095 |
|  |  | **Women** | 1885 | 780 | 1105 | 1.025 | 0.989-1.061 | 0.087 |  |  |  |
|  |  | **Men** | 1875 | 318 | 1557 | 0.976 | 0.932-1.022 | 0.85 |  |  |  |
| **16** | **DGN** | **All** | 941 | 471 | 470 | 1.017 | 0.965-1.071 | 0.268 | 0.992 | 0.886-1.111 | 0.888 |
|  |  | **Women** | 659 | 365 | 294 | 1.02 | 0.958-1.086 | 0.271 |  |  |  |
|  |  | **Men** | 282 | 106 | 176 | 1.01 | 0.919-1.11 | 0.417 |  |  |  |
| **17** | **i2b2-TRD** | **All** | 1879 | 807 | 1072 | 0.978 | 0.941-1.015 | 0.879 | 1.047 | 0.969-1.131 | 0.245 |
|  |  | **Women** | 1076 | 539 | 537 | 0.957 | 0.911-1.006 | 0.958 |  |  |  |
|  |  | **Men** | 803 | 268 | 535 | 1.006 | 0.948-1.068 | 0.419 |  |  |  |
| **18** | **RADIANT-IRISH** | **All** | 448 | 109 | 339 | 0.904 | 0.819-0.997 | 0.978 | 1.217 | 0.954-1.556 | 0.114 |
|  |  | **Women** | 268 | 90 | 178 | 0.869 | 0.776-0.969 | 0.993 |  |  |  |
|  |  | **Men** | 180 | 19 | 161 | 1.057 | 0.851-1.316 | 0.307 |  |  |  |
| **19** | **RADIANT-DEN** | **All** | 649 | 133 | 516 | 1.003 | 0.914-1.101 | 0.471 | 0.946 | 0.781-1.145 | 0.572 |
|  |  | **Women** | 303 | 93 | 210 | 1.025 | 0.91-1.156 | 0.341 |  |  |  |
|  |  | **Men** | 346 | 40 | 306 | 0.97 | 0.835-1.126 | 0.654 |  |  |  |
| **20** | **NESDA/NTR** | **All** | 3097 | 1494 | 1603 | 1.01 | 0.978-1.043 | 0.278 | 1.001 | 0.935-1.07 | 0.986 |
|  |  | **Women** | 1997 | 1021 | 976 | 1.009 | 0.97-1.05 | 0.324 |  |  |  |
|  |  | **Men** | 1100 | 473 | 627 | 1.012 | 0.958-1.07 | 0.33 |  |  |  |
| **21** | **Rotterdam** | **All** | 1286 | 247 | 1039 | 1.001 | 0.947-1.058 | 0.484 | 0.89 | 0.786-1.006 | 0.063 |
|  |  | **Women** | 784 | 184 | 600 | 1.036 | 0.969-1.107 | 0.151 |  |  |  |
|  |  | **Men** | 502 | 63 | 439 | 0.922 | 0.83-1.021 | 0.939 |  |  |  |

**Supplementary Table 4. Forest plot illustrating the odds ratio for depression over increasing T-PRS by PGC cohorts, as well as the overall pooled odds ratio; Meta-analysis including both sexes of 21 PGC cohorts.**

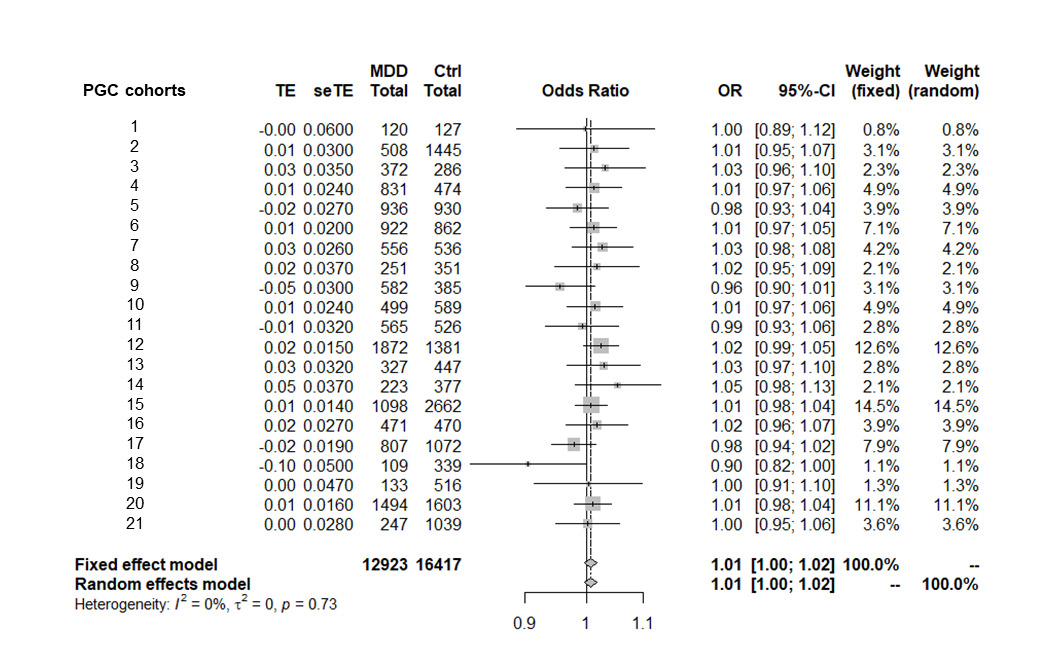

*Abbreviations: TE = Logistic regression coefficient for T-PRS; seTE = standard error of the logistic regression coefficient for T-PRS; MDD Total = Total number of depression cases; Ctrl Total = Total number of controls; OR = odds ratio; 95%-CI = 95% confidence interval; PGC = Psychiatric Genomics Consortium.*

**Supplementary Table 5. Forest plot illustrating the odds ratio for depression over increasing T-PRS by PGC cohorts, as well as the overall pooled odds ratio; Meta-analysis including female subjects only of 21 PGC cohorts.**

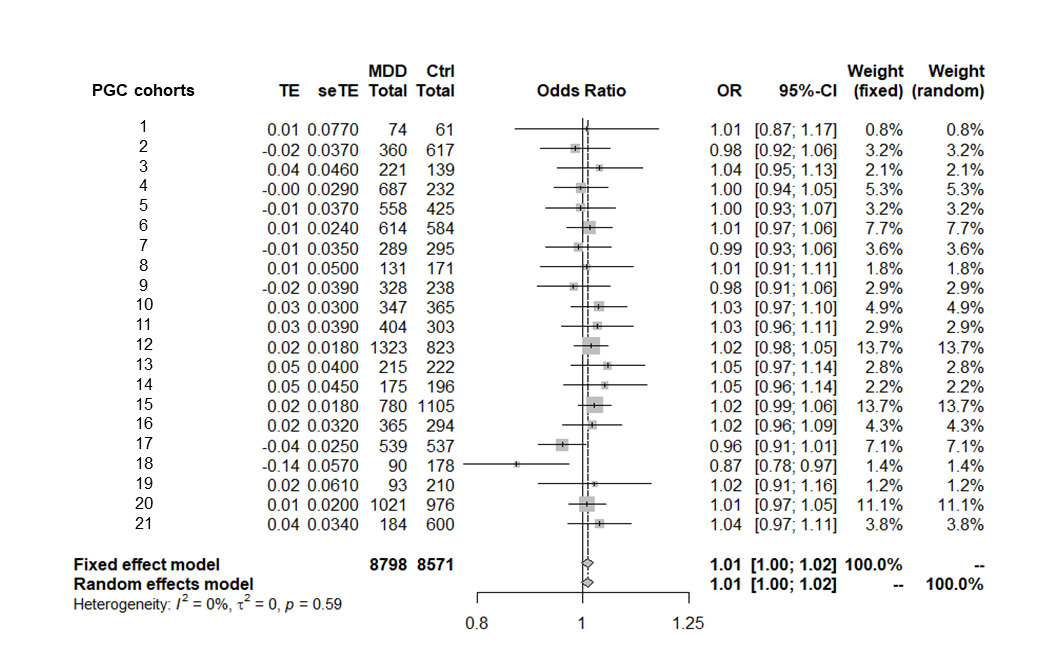

*Abbreviations: TE = Logistic regression coefficient for T-PRS; seTE = standard error of the logistic regression coefficient for T-PRS; MDD Total = Total number of depression cases; Ctrl Total = Total number of controls; OR = odds ratio; 95%-CI = 95% confidence interval; PGC = Psychiatric Genomics Consortium.*

**Supplementary Table 6. Forest plot illustrating the odds ratio for depression over increasing T-PRS by PGC cohorts, as well as the overall pooled odds ratio; Meta-analysis including male subjects only of 21 PGC cohorts.**

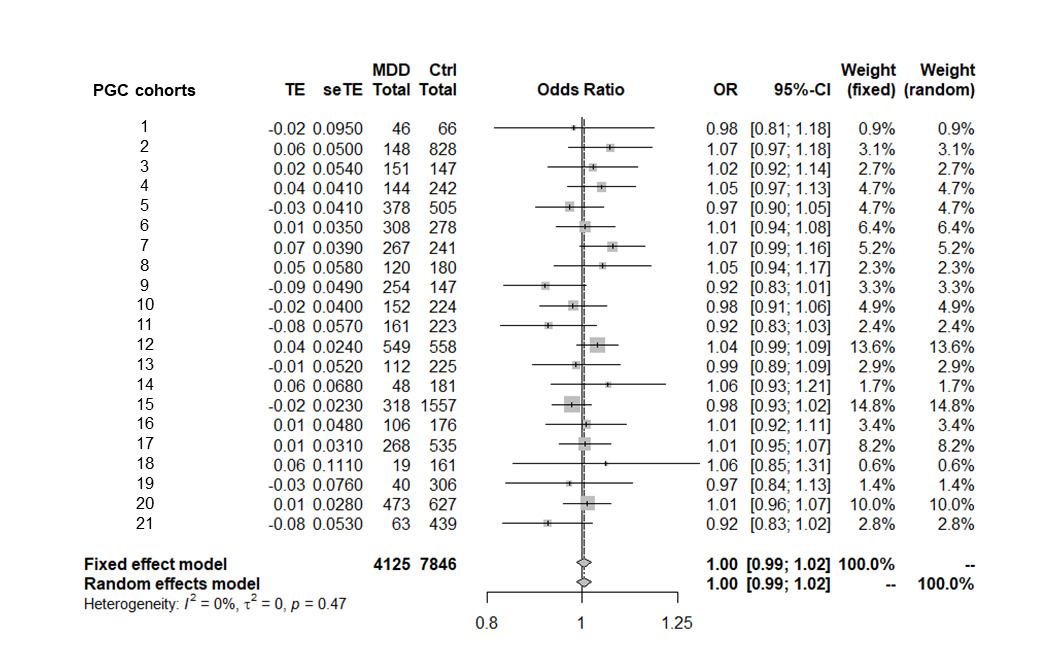

*Abbreviations: TE = Logistic regression coefficient for T-PRS; seTE = standard error of the logistic regression coefficient for T-PRS; MDD Total = Total number of depression cases; Ctrl Total = Total number of controls; OR = odds ratio; 95%-CI = 95% confidence interval; PGC = Psychiatric Genomics Consortium.*

**Supplementary Table 7. Sex-specific main effects of T-PRS on vertex-wise local gyrification**

| **Cluster** | **Max** | **CWP** | **Size (mm^2^)** | **Annot.** |
| --- | --- | --- | --- | --- |
| **Men** |  |  |  |  |
| 1 | -2.970 | .006 | 760 | rostral middle frontal, lh |
| 2 | -3.151 | <.001 | 1965 | lingual, rh |
| 3 | 2.348 | .003 | 806 | superior parietal, rh |
| 4 | 2.109 | .008 | 662 | fusiform, rh |
| **Women** |  |  |  |  |
| 1 | -3.450 | <.001 | 4475 | rostral middle frontal, lh |
| 2 | -3.422 | <.001 | 2734 | superior frontal, lh |
| 3 | -2.118 | .004 | 785 | cuneus, lh |
| 4 | -3.598 | .004 | 782 | fusiform, lh |
| 5 | -2.951 | .014 | 706 | superior temporal, lh |
| 6 | -4.006 | <.001 | 2130 | paracentral, rh |
| 7 | -3.739 | <.001 | 1203 | caudal middle frontal, rh |
| 8 | -2.180 | <.001 | 929 | pars opercularis, rh |
| 9 | -2.880 | .003 | 787 | caudal middle frontal, rh |

*Annot. = Desikan-Killiany atlas label corresponding to the peak vertex; CWP = cluster-wise p-value; Max. = maximum -log10(p) where p is the p-value at the peak vertex*

Clusters in which T-PRS had a significant main effect on vertex-wise local gyrification.

**Supplementary Table 8. Results of post-hoc linear regressions, confirming sex-specific main effects of T-PRS on regional volume or cluster-wise gyrification**

|  | **Tested with psychiatric diagnosis as an additional covariate** | | **Tested in restricted sample, excluding participants with a psychiatric diagnosis** | |
| --- | --- | --- | --- | --- |
| **Region of interest** | **t** | **p** | **t** | **p** |
| **Men** |  |  |  |  |
| Cluster #1, local gyrification | -2.894 | .004 | -2.411 | .017 |
| Cluster #2, local gyrification | -3.387 | .001 | -3.483 | .001 |
| Cluster #3, local gyrification | 2.507 | .013 | 2.074 | .040 |
| Cluster #4, local gyrification | 2.279 | .024 | 1.492 | .138 |
| **Women** |  |  |  |  |
| Amygdala, volume | -3.727 | <.001 | -3.740 | <.001 |
| Cluster #1, local gyrification | -3.338 | .001 | -3.055 | .003 |
| Cluster #2, local gyrification | -3.652 | <.001 | -3.225 | .001 |
| Cluster #3, local gyrification | -2.598 | .010 | -3.025 | .003 |
| Cluster #4, local gyrification | -2.772 | .006 | -2.570 | .011 |
| Cluster #5, local gyrification | -2.893 | .004 | -2.682 | .008 |
| Cluster #6, local gyrification | -3.520 | .001 | -3.368 | .001 |
| Cluster #7, local gyrification | -3.344 | .001 | -3.320 | .001 |
| Cluster #8, local gyrification | -2.715 | .007 | -2.656 | .009 |
| Cluster #9, local gyrification | -3.079 | .002 | -2.934 | .004 |

**Supplementary Table 9. Sex-specific main effects of familial MDD on volume or local gyrification in T-PRS-associated regions or clusters**

| **Brain-based phenotype** | **t** | **p** |
| --- | --- | --- |
| **Men** |  | **pcrit < .0125** |
| Cluster #1, local gyrification | **-2.566** | **.011** |
| Cluster #2, local gyrification | -1.857 | .065 |
| Cluster #3, local gyrification | -0.255 | .799 |
| Cluster #4, local gyrification | -0.250 | .803 |
| **Women** |  | **pcrit < .005** |
| Amygdala, volume | -0.387 | .699 |
| Cluster #1, local gyrification | 1.267 | .206 |
| Cluster #2, local gyrification | -0.081 | .936 |
| Cluster #3, local gyrification | 1.247 | .214 |
| Cluster #4, local gyrification | 0.353 | .724 |
| Cluster #5, local gyrification | -0.221 | .825 |
| Cluster #6, local gyrification | 0.866 | .387 |
| Cluster #7, local gyrification | 0.031 | .975 |
| Cluster #8, local gyrification | -0.248 | .805 |
| Cluster #9, local gyrification | -1.385 | .167 |

*bold indicates significance, as determined using pcrit ≤ .0125 (men), 005 (women)*
